# Associations between serum estradiol and estrone and Alzheimer’s disease biomarkers: an analysis in female participants from the European Prevention of Alzheimer’s Dementia Longitudinal Cohort Study (EPAD LCS)

**DOI:** 10.64898/2026.05.27.26354257

**Authors:** Juehyun Shin, Graciela Muniz-Terrera, Craig W. Ritchie, Jean Manson, Susan Plachecki, Clemens Kirschbaum, Sarah Gregory

## Abstract

**INTRODUCTION:** Postmenopausal estrogen decline may contribute to Alzheimer’s disease (AD) risk, but longitudinal evidence linking circulating estrogens to cerebrospinal fluid (CSF) biomarkers is lacking.

**METHODS:** We analyzed 866 female participants from the European Prevention of AD Longitudinal Cohort Study with baseline serum estradiol and estrone measured by liquid chromatography tandem mass spectrometry and repeated CSF measurements of amyloid-beta (Aβ)_42_, phosphorylated (p) Tau_181_, and total (t) Tau.

**RESULTS:** Neither estradiol nor estrone was associated with longitudinal Aβ_42_. Higher estradiol was associated with lower baseline tau and slower tau increases over time. Baseline estradiol-tau associations were stronger in apolipoprotein E (*APOE*) *ε4* carriers, though *APOEε4* did not modify longitudinal associations. Amyloid positivity did not moderate hormone-tau associations but was associated with steeper tau increases over time. Estrone showed no significant associations.

**DISCUSSION:** These findings suggest a more consistent relationship between estradiol and tau-related rather than amyloid-related pathology.

## 1. BACKGROUND

Globally, dementia prevalence is consistently higher in females than males, beginning in late midlife with the gap widening across older age groups.^1^ Consistent with this epidemiologic pattern, females comprise a larger share of individuals living with Alzheimer’s disease (AD) dementia, and autopsy-based studies suggest females can show higher overall AD neuropathology, particularly greater tau tangle density.^2^ Analyses from the Religious Orders Study and the Memory and Aging Project demonstrated that female participants had greater tau tangle burden than males across multiple brain regions, even after propensity score matching on demographics, apolipoprotein E *ε4* (*APOEε4*) allele count, and other key factors.^3^

Beyond longevity, biological sex differences, particularly sex hormones, have emerged as critical factors that may explain these disparities in AD pathogenesis and risk.^4–7^ Preclinical work suggests that estrogens are neuroactive and may exert neuroprotective effects relevant to AD, including reducing Aβ-related toxicity and modulating tau hyperphosphorylation.^8–11^ The menopausal transition is characterized by a rapid decline in endogenous estrogens, which has been hypothesized to increase vulnerability to AD-relevant pathophysiology in females.^7,9–11^ Observational work further suggests that shorter lifetime endogenous estrogen exposure and earlier menopause may be associated with elevated risk of cognitive impairment and dementia outcomes.^9,12^ Consistent with a menopause-timing framework that emphasizes downstream tau vulnerability, autopsy evidence indicates that earlier menopause can amplify the association between reduced synaptic integrity and tau tangle burden.^12^

While preclinical evidence supports estrogen’s neuroprotective role, translating these findings to human populations has proven challenging. Human studies examining estrogen-AD relationships (including biomarker and cognitive endpoints) and hormone replacement therapy (HRT) have yielded mixed findings. This heterogeneity likely reflects differences in the timing of initiation relative to menopause (including the proposed “critical window”), formulation (e.g., estrogen-only vs. estrogen-progestogen therapy), and baseline characteristics such as genotype and cardiovascular health.^7,9–11,13,14^ A major limitation is reliance on indirect proxies of estrogen exposure (e.g., reproductive history, menopausal status, or HRT use) rather than direct measurement of circulating hormone levels and receptor-mediated signaling activity.^8,9^

Longitudinal studies examining circulating estrogens in relation to cerebrospinal fluid (CSF) AD biomarker trajectories in this population are lacking. The present study addresses this gap by examining associations between serum estrogen levels (estradiol and estrone) and CSF biomarkers of AD pathology in females. We hypothesized that, in females, higher serum estradiol and estrone levels would be associated with slower longitudinal increases in CSF AD biomarkers, amyloid-beta 42 (Aβ_42_), phosphorylated tau 181 (pTau_181_), and total tau (tTau), and that these associations would differ by *APOEε4* carrier status and amyloid positivity.

## 2. METHODS

### 2. 1. Participants

This study utilized data and biological samples from biologically female participants in the European Prevention of AD Longitudinal Cohort Study (EPAD LCS). The EPAD LCS enrolled individuals aged 50+ years representing a spectrum of risk for AD dementia, spanning from healthy volunteers to those with mild cognitive impairment.^15–17^ Participants were excluded if they had missing data on demographic characteristics, *APOEε4* carrier status, estrogen measures (estradiol or estrone), or CSF biomarkers (Aβ_42_, pTau_181_, or tTau). The participant flow diagram (**Figure 1a**) summarizes how the analytic sample was derived from the EPAD LCS cohort. All participants provided informed consent, and local ethical approval was obtained from ethics committees at each research site. All procedures were conducted in accordance with the Helsinki Declaration.

**Figure 1.**
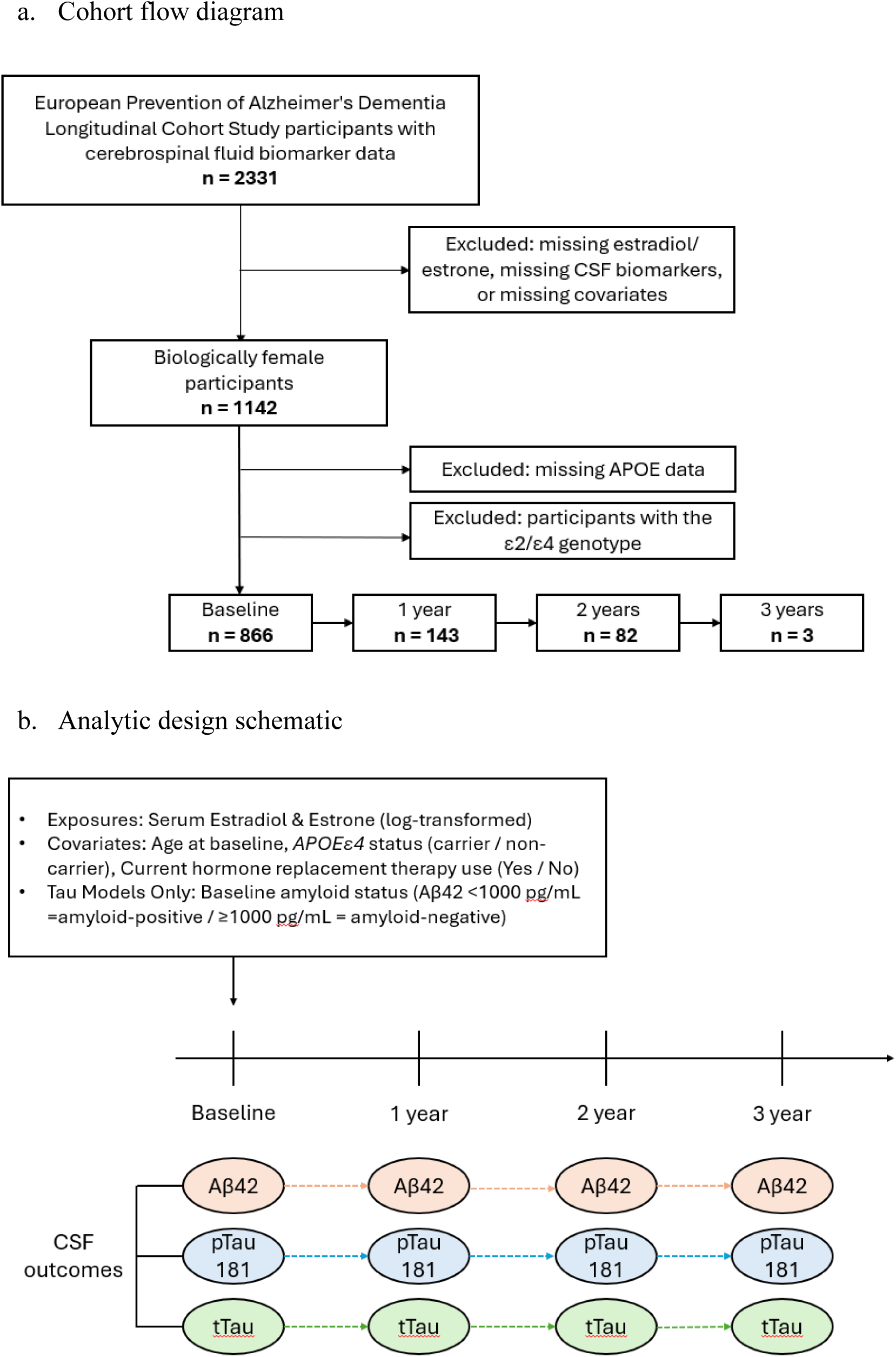
Participant flow diagram and analytic design schematic

**Table 1.**
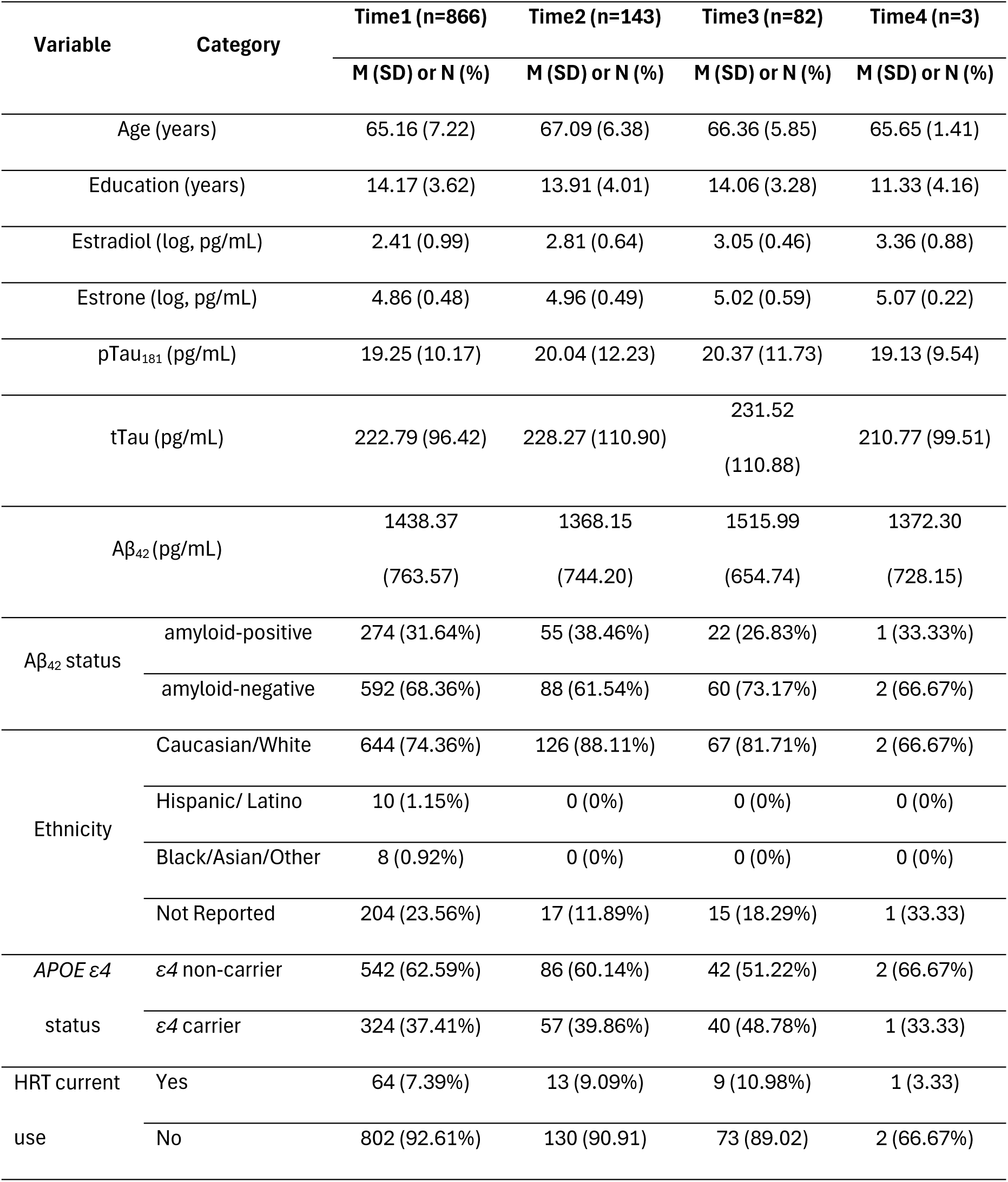
Sample Characteristics.

| Variable | Category | Time1 (n=866) | Time2 (n=143) | Time3 (n=82) | Time4 (n=3) |
| --- | --- | --- | --- | --- | --- |
|  |  | M (SD) or N (%) | M (SD) or N (%) | M (SD) or N (%) | M (SD) or N (%) |
| Age (years) |  | 65.16 (7.22) | 67.09 (6.38) | 66.36 (5.85) | 65.65 (1.41) |
| Education (years) |  | 14.17 (3.62) | 13.91 (4.01) | 14.06 (3.28) | 11.33 (4.16) |
| Estradiol (log, pg/mL) |  | 2.41 (0.99) | 2.81 (0.64) | 3.05 (0.46) | 3.36 (0.88) |
| Estrone (log, pg/mL) |  | 4.86 (0.48) | 4.96 (0.49) | 5.02 (0.59) | 5.07 (0.22) |
| pTau<sub>181</sub> (pg/mL) |  | 19.25 (10.17) | 20.04 (12.23) | 20.37 (11.73) | 19.13 (9.54) |
| tTau (pg/mL) |  | 222.79 (96.42) | 228.27 (110.90) | 231.52<br>(110.88) | 210.77 (99.51) |
| Aβ<sub>42</sub> (pg/mL) | | 1438.37<br>(763.57) | 1368.15<br>(744.20) | 1515.99<br>(654.74) | 1372.30<br>(728.15) |
| Aβ<sub>42</sub> status | amyloid-positive | 274 (31.64%) | 55 (38.46%) | 22 (26.83%) | 1 (33.33%) |
|  | amyloid-negative | 592 (68.36%) | 88 (61.54%) | 60 (73.17%) | 2 (66.67%) |
| Ethnicity | Caucasian/White | 644 (74.36%) | 126 (88.11%) | 67 (81.71%) | 2 (66.67%) |
|  | Hispanic/ Latino | 10 (1.15%) | 0 (0%) | 0 (0%) | 0 (0%) |
|  | Black/Asian/Other | 8 (0.92%) | 0 (0%) | 0 (0%) | 0 (0%) |
|  | Not Reported | 204 (23.56%) | 17 (11.89%) | 15 (18.29%) | 1 (33.33) |
| APOE ε4 | ε4 non-carrier | 542 (62.59%) | 86 (60.14%) | 42 (51.22%) | 2 (66.67%) |
| status | ε4 carrier | 324 (37.41%) | 57 (39.86%) | 40 (48.78%) | 1 (33.33) |
| HRT current | Yes | 64 (7.39%) | 13 (9.09%) | 9 (10.98%) | 1 (3.33) |
| use | No | 802 (92.61%) | 130 (90.91) | 73 (89.02) | 2 (66.67%) |

### 2. 2. Estradiol and estrone measurement

Blood samples collected as part of the EPAD LCS were analyzed for estradiol and estrone using liquid chromatography tandem mass spectrometry (LC-MS/MS); details of the online solid-phase extraction–high-performance liquid chromatography (SPE HPLC) method have been reported previously.^18^ In contrast to the instrumentation and mobile phases described, the analysis was performed on a SCIEX 6500+ triple quad mass spectrometer.

The present method used 100% methanol as mobile phase A, 0.2 mM ammonium fluoride in water as mobile phase B, C-A was water (1st elution step and re-equilibration step), and C-B was 100% LC-MS grade methanol (washing columns and needle). The flow rate was maintained at 3 mL/minute, with a column temperature of 40°C, an injection volume of 50 μL, with a total run time of 9 minutes. The LC eluent was introduced to the MS through the turbo-ion-spray probe in negative ionization mode for estrogens, with multiple reaction monitoring (MRM) used. The following ion source parameters were used: curtain gas (CUR) set to 35, collision gas (CAD) was set to 7, ion spray voltage (IS) was -4500 V, ion source temperature (TEM) was 350°C, and ion source gas 1 and ion source gas 2 were both set to 65.

A quantifier and a qualifier ion were detected for each analyte in each sample.

Established calibration curves were used for quality control, alongside quality control samples included in the runs with known concentrations of analytes of interest to check for accuracy of detection. Circulating estrogen levels (estradiol and estrone) were measured as continuous variables. Estriol was also analyzed, but as only a few participants had detectable concentrations, no analysis was carried out for this hormone.

### 2. 3. CSF biomarker assessment

CSF samples were analyzed at the Clinical Neurochemistry Laboratory, University of Gothenburg, Sweden, using the Roche Diagnostics Elecsys platform for Alzheimer’s disease biomarkers, including Aβ_42_, pTau_181_, and tTau. The established cut-offs for the EPAD LCS are defined as: Aβ_42_ <1000 pg/mL (amyloid-positive).^19^ For data processing, values below the reporting limits were set to the limit (pTau_181_ = 8 pg/mL, tTau = 80 pg/mL, Aβ_42_ = 200 pg/mL), and Aβ_42_ values >1700 pg/mL were handled using recalculated results.

### 2. 4. *APOE* genotyping

*APOEε4* data were categorized into two groups: a non-carrier group (including *ε2/ε2*, *ε2/ε3*, and *ε3/ε3* genotypes) and a carrier group (including *ε3/ε4* and *ε4/ε4* genotypes). Participants with the *ε2/ε4* genotype were excluded from the analysis as this genotype includes both protective and risk alleles.^20^

### 2. 5. Hormone replacement therapy status

HRT information was derived from recorded HRT medication use and included both estrogen-only and combined estrogen plus progestogen preparations. Estrogen-only formulations included estradiol (including estradiol hemihydrate and estradiol valerate), estriol, and estrone preparations, as well as transdermal estradiol patch products (e.g., FemSeven and Oestranorm). Combined regimens included estradiol plus progestogen products such as Estalis SequiDot (estradiol + norethisterone), Fem7 Combi (estradiol + levonorgestrel), Kliogest (estradiol + norethisterone acetate), and Zumeston (estradiol + norethisterone acetate). Preparations encompassed both oral and transdermal routes and a range of doses. For the primary analyses, HRT status was coded as a binary indicator of current use at baseline (yes/no).

### 2. 6. Statistical Analysis

Baseline characteristics were summarized using means (SD) for continuous variables and frequencies (%) for categorical variables, and estradiol and estrone were natural log-transformed to improve model fit and reduce the influence of extreme values. Longitudinal associations between baseline serum estrogen levels and repeated CSF biomarkers were examined using linear mixed-effects models with random interceptions and random slopes for time. Age at visit was used as the time metric and mean-centered by subtracting the overall mean age across all observations, 65.50 years. Thus, model intercepts correspond to expected biomarker levels at age 65.50 years. Time interaction terms were included for all covariates. Models adjusted for baseline age, *APOEε4* carrier status, and current HRT use, and *APOEε4* carrier moderation was evaluated; tau models additionally included baseline amyloid status as covariate and tested amyloid moderation. **Figure 1b** provides a schematic overview of the analytic design, including timing of baseline hormone measurement and repeated CSF biomarker assessments, and the covariates included in the mixed-effects models. To account for multiple comparisons, we applied the Benjamini and Hochberg^21^ false discovery rate procedure to the primary hormone-related terms within each biomarker-specific primary model. Terms with BH-adjusted q-values < 0.05 were considered statistically significant. As an exploratory follow-up to interaction testing, we conducted stratified mixed-effects models by APOE *ε4* carrier status to facilitate interpretation of hormone associations within strata. Given substantial attrition after the first visit, we compared baseline characteristics between participants with any follow-up and those with baseline-only observations to assess potential selection due to differential follow-up. As a sensitivity analysis addressing potential bias due to differential follow-up, we repeated the primary mixed-effects models restricted to participants with at least one follow-up visit.

Moderation terms, sensitivity analyses, and exploratory stratified analyses are shown with nominal p-values. Analyses were conducted using Stata (version 19).

## 3. RESULTS

After exclusion of 21 participants with the *ε2/ε4* genotype, the analytic sample comprised 866 participants. At baseline, mean age was 65.2 years (SD 7.2). Mean log-transformed estradiol and estrone concentrations were 2.41 (SD 0.99) and 4.86 (SD 0.48), respectively (**Table1**). Mean CSF pTau_181_, tTau, and Aβ_42_ concentrations were 19.25 pg/mL (SD 10.17), 222.79 pg/mL (SD 96.42), and 1438.37 pg/mL (SD 763.57), respectively. Overall, 31.6% of participants were amyloid-positive, 37.4% were *APOE*ε4 carriers, and 7.4% reported current HRT use. Sample size declined across follow-up visits to 143 at Time 2, 82 at Time 3, and 3 at Time 4.

In attrition analyses (**Table S1** in **Supplement)**, participants with any follow-up data (n = 189) had higher baseline log-transformed estradiol and estrone concentrations and lower baseline CSF Aβ_42_ concentrations than those with baseline-only data (n = 677) (all *p* ≤ 0.05). Baseline age, education, pTau_181_, tTau, amyloid positivity, *APOEε4* carrier status, and current HRT use did not differ by follow-up status. Ethnicity differed between groups (*p* < 0.001), with participants contributing follow-up data more likely to be categorized as White and less likely to have ethnicity recorded as not reported.

### 3.1. Estradiol / Estrone and Aβ_42_

Neither log-transformed estradiol nor estrone was associated with baseline Aβ_42_ or with change in Aβ_42_ over time (**Table 2**). Older baseline age was associated with lower Aβ_42_ concentrations in both the estradiol and estrone models (both *p* < 0.001). *APOEε4* carrier status was likewise associated with lower baseline Aβ_42_ in the primary models. In models including *APOEε4* interaction terms, there was no evidence that *APOEε4* modified the associations of estradiol or estrone with baseline Aβ_42_ or its change over time. Current HRT use was not associated with baseline Aβ_42_ or its change

**Table 2.** Linear mixed model estimates of CSF Aβ_42_ in relation to serum estradiol and estrone.

| Predictor | Model 1 |  |  |  | Model 2 |  |  |  |
| --- | --- | --- | --- | --- | --- | --- | --- | --- |
|  | Baseline |  | Slope |  | Baseline |  | Slope |  |
| | $\beta$ (SE) | *P* | $\beta$ (SE) | *P* | $\beta$ (SE) | *P* | $\beta$ (SE) | *P* |
| Estradiol |  |  |  |  |  |  |  |  |
| Intercept | 6114.47<br>(908.37) | <0.001*** | 97.94<br>(35.87) | 0.006** | 6102.47<br>(909.27) | <0.001*** | 101.31<br>(36.65) | 0.006** |
| Age at baseline<br>(years) | -67.66 (13.76) | <0.001*** | -0.58 (0.46) | 0.21 | -67.62 (13.76) | <0.001*** | -0.60 (0.47) | 0.20 |
| APOE $\epsilon 4$ carrier<br>(vs non-carrier) | -400.28 (52.25) | <0.001*** | -20.85<br>(7.38) | 0.005** | -374.11<br>(137.95) | 0.007** | -26.65<br>(18.26) | 0.14 |
| Current HRT<br>use (vs no) | 102.51 (99.63) | 0.30 | -2.49<br>(14.42) | 0.86 | 102.72<br>(100.03) | 0.30 | -2.30 (14.44) | 0.87 |
| ln (Estradiol) | -29.87 (27.34) | 0.27 | 3.66 (3.40) | 0.28 | -25.77 (35.38) | 0.47 | 2.77 (4.38) | 0.53 |
| APOE $\epsilon 4$ carrier<br>× ln (Estradiol) | . | . | . | . | -10.60 (53.79) | 0.84 | 2.23 (6.68) | 0.74 |
| Estrone |  |  |  |  |  |  |  |  |
| Intercept | 5883.45<br>(952.35) | <0.001*** | 86.89<br>(51.38) | 0.09 | 5883.91<br>(976.15) | <0.001*** | 80.82<br>(58.79) | 0.17 |
| Age at baseline<br>(years) | -67.61 (13.70) | <0.001*** | -0.71 (0.46) | 0.12 | -67.57 (13.71) | <0.001*** | -0.71 (0.46) | 0.13 |
| APOE $\epsilon 4$ carrier<br>(vs non-carrier) | -398.19 (52.31) | <0.001*** | -21.14<br>(7.37) | 0.004** | -398.30<br>(546.51) | 0.47 | -7.41 (69.63) | 0.92 |
| Current HRT<br>use (vs no) | 58.08 (101.92) | 0.57 | -3.82<br>(14.73) | 0.80 | 58.92 (102.24) | 0.56 | -3.93 (14.77) | 0.79 |
| ln (Estrone) | 33.88 (58.13) | 0.56 | 5.97 (7.24) | 0.41 | 33.15 (74.14) | 0.66 | 7.15 (9.24) | 0.44 |
| APOE $\epsilon 4$ carrier<br>× ln (Estrone) | . | . | . | . | -0.11 (112.45) | 1.00 | -2.79 (14.17) | 0.84 |

Notes: \* $p < .05$ . \*\* $p < .01$ . \*\*\* $p < .001$ . † Survives FDR correction at $q < .05$

### 3.2. Estradiol and Tau

In models of pTau_181_ (**Table 3**), higher circulating estradiol was associated with lower baseline pTau_181_ and a slower increase in pTau_181_ over time in the primary model (Model 1: both primary estradiol terms survived BH-FDR correction; baseline: β = -1.00, *p* = 0.004, BH-FDR *q* < 0.05; slope: β = -0.07, *p* = 0.047, BH-FDR *q* < 0.05). *APOEε4* carrier status was associated with higher baseline pTau_181_ and a steeper increase. In models including interaction terms, the cross-sectional association between estradiol and pTau_181_ differed by *APOEε4* status, whereas there was no evidence that *APOEε4* modified the association with change over time (**Figure 2a**, **2b**). There was likewise no evidence that amyloid status modified the cross-sectional or longitudinal associations of estradiol with pTau_181_. Across models, amyloid positivity was associated with a steeper increase in pTau_181_.

**Figure 2.**
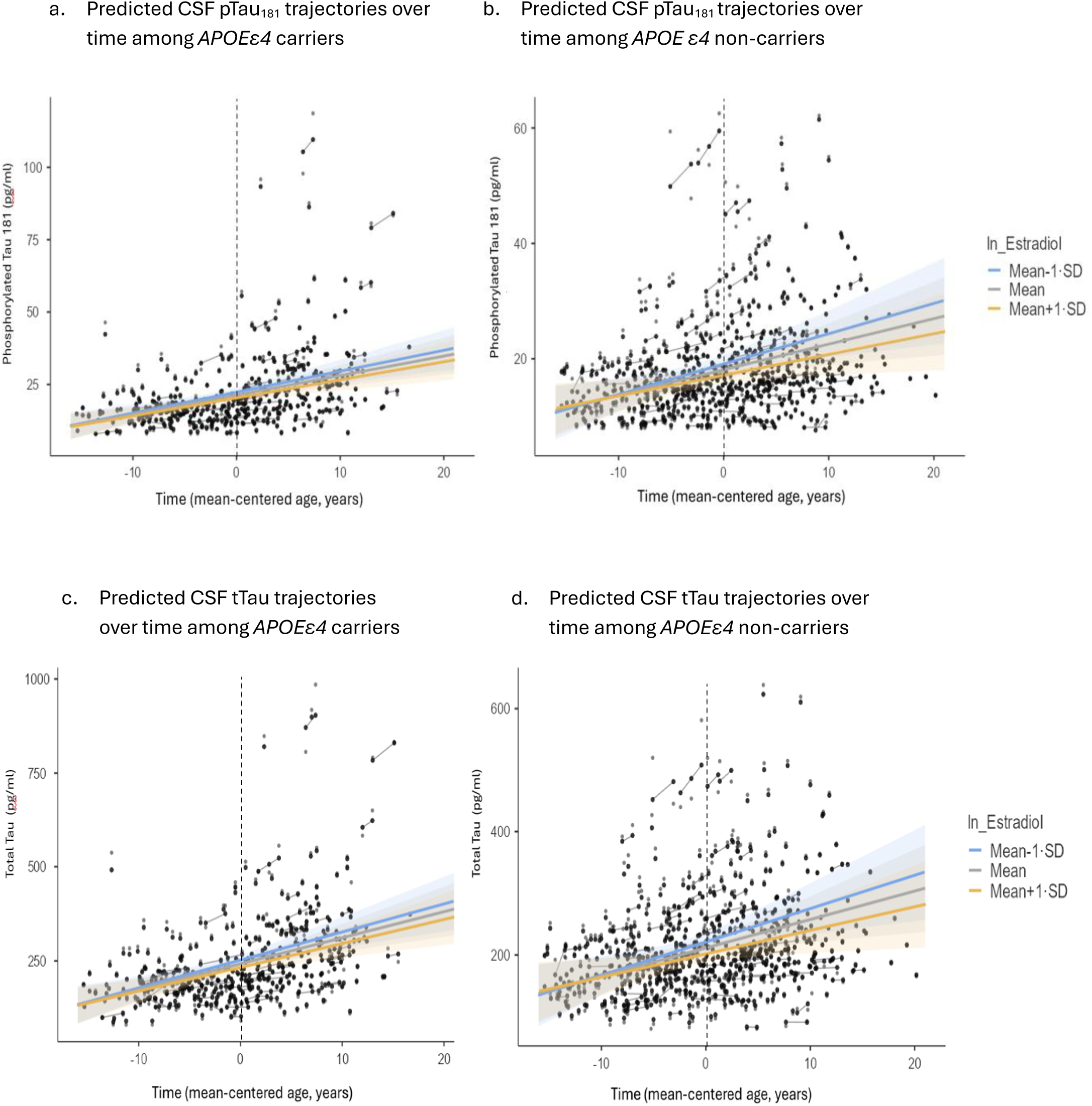
Model-predicted CSF tau trajectories over follow-up by baseline estradiol (low vs high) and APOE ε4 carrier status. Predicted values were derived from linear mixed-effects models using mean-centered age at each visit as the time scale. Estradiol strata represent low versus high baseline concentrations (mean-1 SD vs mean+1 SD). Shaded bands indicate 95% confidence intervals.

**Table 3.** Linear mixed model estimates of CSF pTau_181_ in relation to serum estradiol.

| Predictor | Model 1 |  |  |  | Model 2 |  |  |  | Model 3 |  |  |  |
| --- | --- | --- | --- | --- | --- | --- | --- | --- | --- | --- | --- | --- |
|  | Baseline |  | Slope |  | Baseline |  | Slope |  | Baseline |  | Slope |  |
| | $\beta$<br>(SE) | *P* | $\beta$ (SE) | *P* | $\beta$<br>(SE) | *P* | $\beta$ (SE) | *P* | $\beta$ (SE) | *P* | $\beta$ (SE) | *P* |
| Intercept | 18.28<br>(8.81) | 0.038* | -0.16<br>(0.39) | 0.69 | 17.30<br>(8.82) | 0.05 | -0.16<br>(0.40) | 0.68 | 18.05<br>(8.81) | 0.040* | -0.19<br>(0.39) | 0.63 |
| Age at baseline<br>(years) | 0.02<br>(0.13) | 0.86 | 0.01<br>(0.01) | 0.06 | 0.02<br>(0.13) | 0.90 | 0.01<br>(0.01) | 0.09 | 0.02<br>(0.13) | 0.90 | 0.01<br>(0.01) | 0.07 |
| APOE $\epsilon 4$ carrier<br>(vs non-carrier) | 3.27<br>(0.67) | <0.001*** | 0.23<br>(0.08) | 0.004** | 6.51<br>(1.72) | <0.001*** | 0.36<br>(0.20) | 0.07 | 3.19<br>(0.67) | <0.001*** | 0.23<br>(0.08) | 0.005** |
| Current HRT use<br>(vs no) | 1.60<br>(1.24) | 0.20 | 0.04<br>(0.15) | 0.79 | 1.81<br>(1.24) | 0.15 | 0.07<br>(0.15) | 0.67 | 1.65<br>(1.24) | 0.18 | 0.05<br>(0.15) | 0.76 |
| Amyloid-positive<br>(vs negative) | 0.09<br>(0.51) | 0.86 | 0.27<br>(0.07) | <0.001*** | 0.08<br>(0.51) | 0.87 | 0.27<br>(0.07) | <0.001*** | 1.82<br>(1.80) | 0.31 | 0.44<br>(0.20) | 0.030* |
| ln (Estradiol) | -1.00<br>(0.34) | 0.004† | -0.07<br>(0.04) | 0.047† | -0.43<br>(0.44) | 0.34 | -0.05<br>(0.05) | 0.33 | -0.76<br>(0.41) | 0.06 | -0.05<br>(0.04) | 0.26 |
| APOE $\epsilon 4$ carrier<br>× ln (Estradiol) | . | . | . | . | -1.37<br>(0.67) | 0.041* | -0.06<br>(0.07) | 0.40 | . | . | . | . |
| Amyloid-positive<br>× ln (Estradiol) | . | . | . | . | . | . | . | . | -0.66<br>(0.65) | 0.32 | -0.06<br>(0.07) | 0.36 |

Notes: \* $p < .05$ . \*\* $p < .01$ . \*\*\* $p < .001$ . † Survives FDR correction at $q < .05$

In models of tTau (**Table 4**), higher circulating estradiol was associated with lower baseline tTau and a slower increase in tTau over time in the primary model (Model 1; baseline: β = -9.89, *p* = 0.003, BH-FDR *q* < 0.05; slope: β = -0.72, *p* = 0.049, BH-FDR *q* < 0.05). *APOEε4* carrier status was associated with higher baseline tTau and a steeper increase. In models including interaction terms, the cross-sectional association between estradiol and tTau differed by *APOEε4* status, whereas there was no evidence that *APOEε4* modified the association with change over time (**Figure 2c**, **2d**). There was likewise no evidence that amyloid status modified the cross-sectional or longitudinal associations of estradiol with tTau. Across models, amyloid positivity was associated with a steeper increase in tTau over time.

**Table 4.**
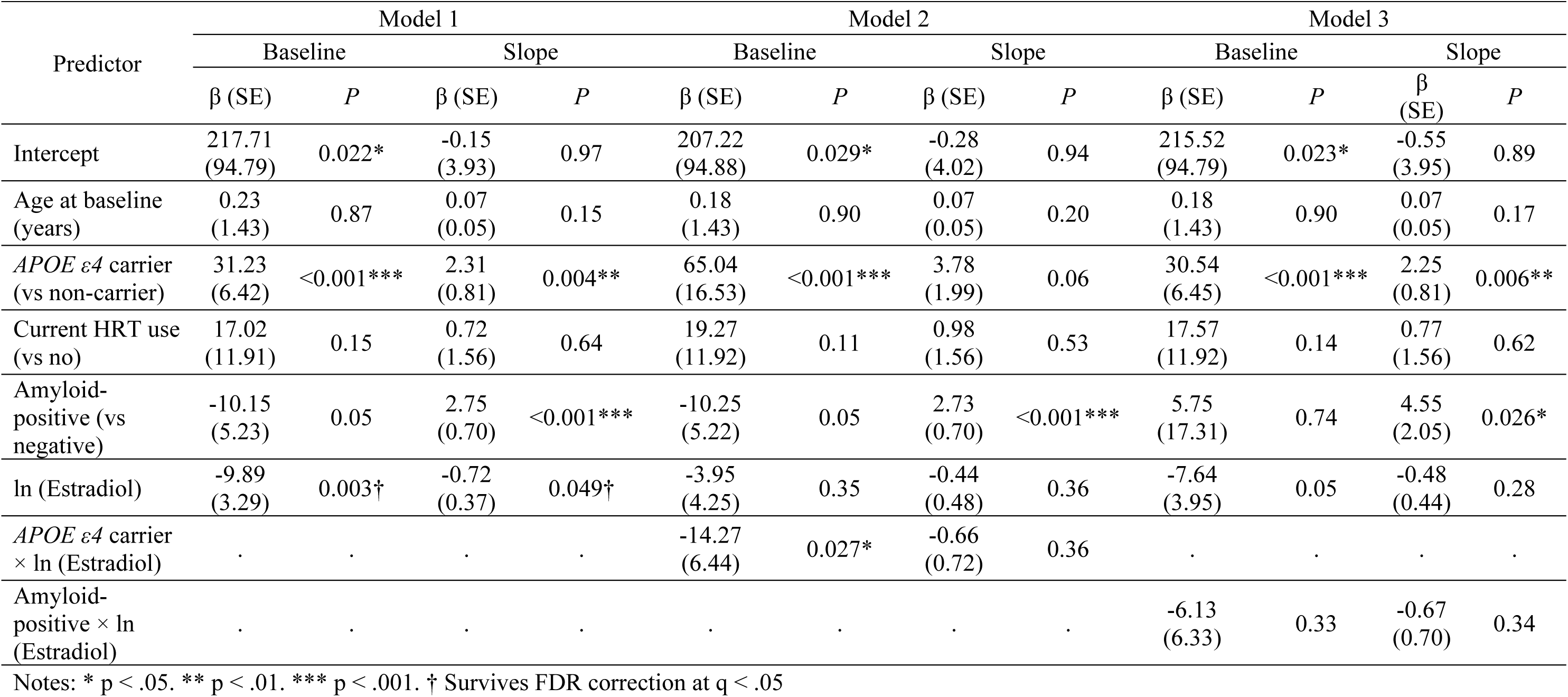
Linear mixed model estimates of CSF tTau in relation to serum estradiol.

### 3.3. Estrone and Tau

In models of pTau_181_ (**Table 5**), circulating estrone was not associated with baseline pTau_181_ or with change in pTau_181_ in the primary model, and no estrone main effects survived BH-FDR correction. *APOEε4* carrier status was associated with higher baseline pTau_181_ and a steeper increase. In models including interaction terms, there was no evidence that *APOEε4* modified the cross-sectional or longitudinal associations of estrone with pTau_181_. There was likewise no evidence that amyloid status modified the cross-sectional or longitudinal associations of estrone with pTau_181_. Across models, amyloid positivity was associated with a steeper increase in pTau_181_.

**Table 5.** Linear mixed model estimates of CSF pTau_181_ in relation to serum estrone.

| Predictor | Model 1 |  |  |  | Model 2 |  |  |  | Model 3 |  |  |  |
| --- | --- | --- | --- | --- | --- | --- | --- | --- | --- | --- | --- | --- |
|  | Baseline |  | Slope |  | Baseline |  | Slope |  | Baseline |  | Slope |  |
| | $\beta$ (SE) | *P* | $\beta$ (SE) | *P* | $\beta$ (SE) | *P* | $\beta$ (SE) | *P* | $\beta$ (SE) | *P* | $\beta$ (SE) | *P* |
| Intercept | 20.72<br>(9.49) | 0.029* | 0.15<br>(0.56) | 0.79 | 17.18<br>(9.84) | 0.08 | 0.05<br>(0.65) | 0.94 | 19.18<br>(9.70) | 0.048* | -0.12<br>(0.59) | 0.85 |
| Age at baseline<br>(years) | 0.04<br>(0.13) | 0.74 | 0.01<br>(0.01) | 0.06 | 0.04<br>(0.13) | 0.77 | 0.01<br>(0.01) | 0.08 | 0.04<br>(0.13) | 0.79 | 0.01<br>(0.01) | 0.08 |
| *APOE* ε4 carrier<br>(vs non-carrier) | 3.30<br>(0.67) | <0.001*** | 0.24<br>(0.08) | 0.003** | 12.65<br>(6.85) | 0.07 | 0.64<br>(0.75) | 0.39 | 3.26<br>(0.67) | <0.001*** | 0.23<br>(0.08) | 0.004** |
| Current HRT use<br>(vs no) | 1.41<br>(1.27) | 0.27 | 0.03<br>(0.16) | 0.83 | 1.53<br>(1.28) | 0.23 | 0.05<br>(0.16) | 0.75 | 1.47<br>(1.27) | 0.25 | 0.02<br>(0.16) | 0.88 |
| Amyloid-positive<br>(vs negative) | 0.06<br>(0.51) | 0.91 | 0.27<br>(0.07) | <0.001*** | 0.04<br>(0.51) | 0.93 | 0.27<br>(0.07) | <0.001*** | 6.88<br>(7.26) | 0.34 | 1.39<br>(0.76) | 0.07 |
| ln (Estrone) | -1.26<br>(0.73) | 0.09 | -0.10<br>(0.08) | 0.20 | -0.46<br>(0.93) | 0.62 | -0.07<br>(0.10) | 0.48 | -0.84<br>(0.84) | 0.32 | -0.04<br>(0.09) | 0.66 |
| *APOE* ε4 carrier<br>× ln (Estrone) | . | . | . | . | -1.93<br>(1.41) | 0.17 | -0.09<br>(0.15) | 0.57 | . | . | . | . |
| Amyloid-positive<br>× ln (Estrone) | . | . | . | . | . | . | . | . | -1.41<br>(1.49) | 0.34 | -0.23<br>(0.15) | 0.14 |

Notes: \* $p < .05$ . \*\* $p < .01$ . \*\*\* $p < .001$ . † Survives FDR correction at $q < .05$

In models of tTau (**Table 6**), circulating estrone was not associated with baseline tTau or with its change in the primary model, and no estrone main effects survived BH-FDR correction. *APOEε4* carrier status was associated with higher baseline tTau and a steeper increase. In models including interaction terms, there was no evidence that *APOEε4* modified the cross-sectional or longitudinal associations of estrone with tTau. There was likewise no evidence that amyloid status modified the cross-sectional or longitudinal associations of estrone with tTau. Across models, amyloid positivity was associated with a steeper increase in tTau.

**Table 6.**
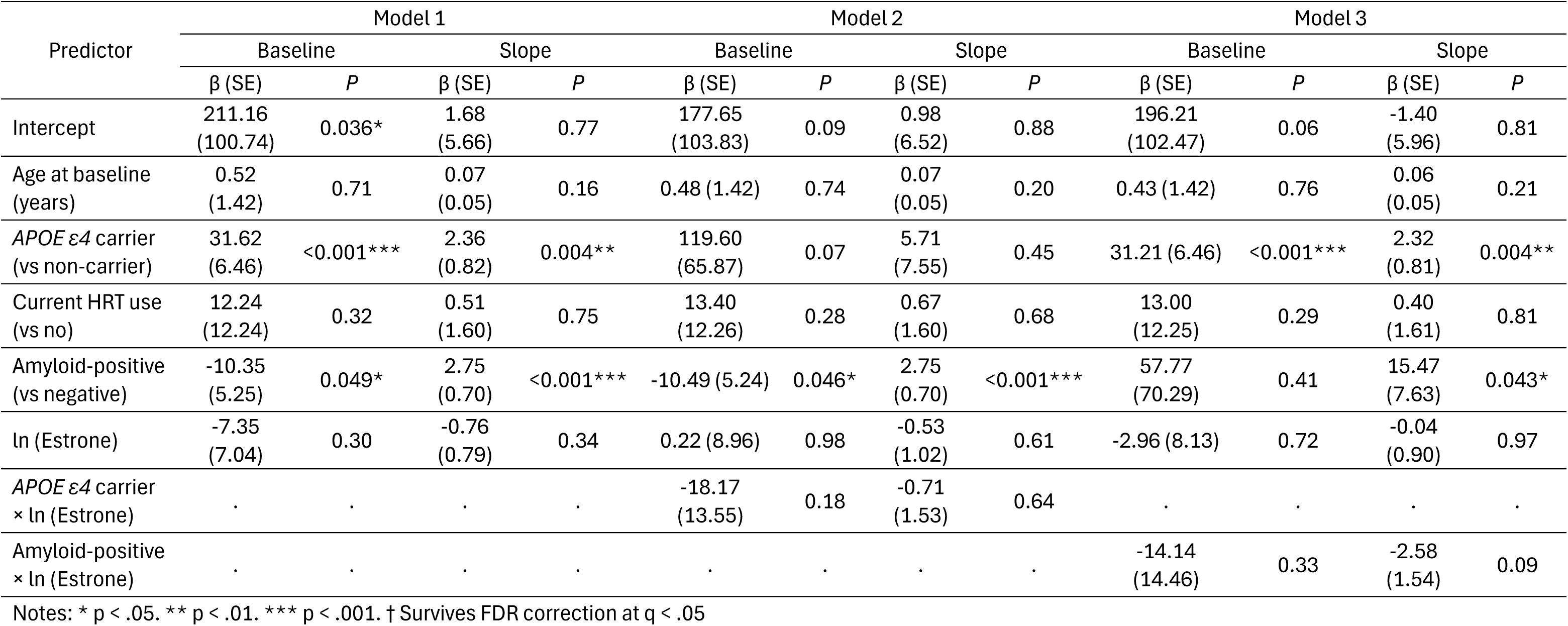
Linear mixed model estimates of CSF tTau in relation to serum estrone.

| Predictor | Model 1 |  |  |  | Model 2 |  |  |  | Model 3 |  |  |  |
| --- | --- | --- | --- | --- | --- | --- | --- | --- | --- | --- | --- | --- |
|  | Baseline |  | Slope |  | Baseline |  | Slope |  | Baseline |  | Slope |  |
| | $\beta$ (SE) | *P* | $\beta$ (SE) | *P* | $\beta$ (SE) | *P* | $\beta$ (SE) | *P* | $\beta$ (SE) | *P* | $\beta$ (SE) | *P* |
| Intercept | 211.16<br>(100.74) | 0.036* | 1.68<br>(5.66) | 0.77 | 177.65<br>(103.83) | 0.09 | 0.98<br>(6.52) | 0.88 | 196.21<br>(102.47) | 0.06 | -1.40<br>(5.96) | 0.81 |
| Age at baseline<br>(years) | 0.52<br>(1.42) | 0.71 | 0.07<br>(0.05) | 0.16 | 0.48 (1.42) | 0.74 | 0.07<br>(0.05) | 0.20 | 0.43 (1.42) | 0.76 | 0.06<br>(0.05) | 0.21 |
| *APOE* ε4 carrier<br>(vs non-carrier) | 31.62<br>(6.46) | <0.001*** | 2.36<br>(0.82) | 0.004** | 119.60<br>(65.87) | 0.07 | 5.71<br>(7.55) | 0.45 | 31.21 (6.46) | <0.001*** | 2.32<br>(0.81) | 0.004** |
| Current HRT use<br>(vs no) | 12.24<br>(12.24) | 0.32 | 0.51<br>(1.60) | 0.75 | 13.40<br>(12.26) | 0.28 | 0.67<br>(1.60) | 0.68 | 13.00<br>(12.25) | 0.29 | 0.40<br>(1.61) | 0.81 |
| Amyloid-positive<br>(vs negative) | -10.35<br>(5.25) | 0.049* | 2.75<br>(0.70) | <0.001*** | -10.49 (5.24) | 0.046* | 2.75<br>(0.70) | <0.001*** | 57.77<br>(70.29) | 0.41 | 15.47<br>(7.63) | 0.043* |
| ln (Estrone) | -7.35<br>(7.04) | 0.30 | -0.76<br>(0.79) | 0.34 | 0.22 (8.96) | 0.98 | -0.53<br>(1.02) | 0.61 | -2.96 (8.13) | 0.72 | -0.04<br>(0.90) | 0.97 |
| *APOE* ε4 carrier<br>× ln (Estrone) | . | . | . | . | -18.17<br>(13.55) | 0.18 | -0.71<br>(1.53) | 0.64 | . | . | . | . |
| Amyloid-positive<br>× ln (Estrone) | . | . | . | . | . | . | . | . | -14.14<br>(14.46) | 0.33 | -2.58<br>(1.54) | 0.09 |

Notes: \* $p < .05$ . \*\* $p < .01$ . \*\*\* $p < .001$ . † Survives FDR correction at $q < .05$

### 3.4. Sensitivity analyses

Sensitivity analyses restricted to participants with at least one follow-up visit (**Tables S2-S6** in **Supplement**) were broadly consistent with the primary findings. Older baseline age remained associated with lower Aβ_42_ in both estradiol and estrone models, and neither hormone was associated with change in Aβ_42_ (**Table S2**). There was likewise no evidence that *APOEε4* modified the associations of estradiol or estrone with Aβ_42_. In these follow-up-restricted Aβ_42_ models, estrone was positively associated with baseline Aβ_42_, a pattern not observed in the full sample; given the reduced sample and absence of a corresponding estrone-by-time association, this finding should be interpreted cautiously.

For tau biomarkers, the inverse associations of estradiol with pTau_181_ and tTau observed in the primary analyses were attenuated and no longer statistically significant in follow-up-restricted models, with no evidence of effect modification by *APOEε4* or amyloid status (**Tables S3-S4**). Estrone models also remained null for pTau_181_ and tTau (**Tables S5-S6**). Although a nominal estrone-by-time association for pTau_181_ in Model 1 (**Table S5**) and an amyloid × estrone × time interaction for tTau in Model 3 (**Table S6**) were observed, these findings were observed only in the follow-up-restricted analyses and not in the main full-sample models. Given evidence of informative attrition, the substantially smaller restricted sample, and the nominal nature of these associations, these findings should be interpreted cautiously.

### 3.5. Stratified analysis by APOE ε4 carrier status

In stratified models by *APOEε4* carrier status (**Table S7** in **Supplement**), higher circulating estradiol was associated with lower baseline pTau_181_ and lower baseline tTau among *APOEε4* carriers (pTau_181_: β = -1.69, *p* = 0.007; tTau: β = -17.96, *p* = 0.002), whereas corresponding associations were not observed among non-carriers. Estradiol was not associated with longitudinal change in pTau_181_ or tTau in either stratum. These findings were consistent with the primary interaction models, which showed effect modification by *APOEε4* status for the cross-sectional associations of estradiol with pTau_181_ and tTau, but not for change over time. In contrast, estrone was not associated with baseline levels or change in pTau_181_ or tTau in either *APOEε4* stratum, consistent with the null *APOEε4* interaction terms in the primary estrone models. Across stratified models, amyloid positivity was associated with steeper increases in tau biomarkers, particularly among *APOEε4* non-carriers.

## 4. Discussion

Among female participants in the EPAD LCS, baseline estradiol and estrone were not associated with baseline Aβ_42_ concentrations or its change over time, suggesting limited evidence that peripheral estrogen levels relate to amyloid biomarker dynamics in this cohort. In contrast, higher baseline estradiol was associated with lower baseline tau biomarkers (pTau_181_ and tTau) and with modestly slower increases in tau over time in primary models, with cross-sectional estradiol-tau associations differing by *APOEε4* carrier status. Amyloid positivity did not moderate hormone-tau associations but was associated with steeper tau increases over time.

Estrone showed largely non-significant associations with tau outcomes. These associations were attenuated in follow-up-restricted analyses.

The observed inverse associations between circulating estradiol and tau biomarkers are consistent with experimental evidence that estradiol may protect against tau pathology.

Mechanistically, estradiol acts through Estrogen receptor (ER) α, ERβ, and G protein–coupled estrogen receptor (GPER) in neurons and glia, influencing pathways involved in synaptic plasticity, neuronal survival, kinase regulation, oxidative stress, and neuroinflammation.^22–27^ These receptor-mediated effects include AMPK-related signaling and reduced Glycogen Synthase Kinase (GSK) -3β activity, both of which are relevant to tau phosphorylation and cellular homeostasis.^28,29^ Recent work also suggests that estradiol may promote tau clearance and restrain inflammatory processes that accelerate tau accumulation. In tauopathy models, estrogen receptor signaling has been linked to enhanced autophagy and improved tau proteostasis, including through GPER-related mechanisms.^30^ Estradiol has also been shown to suppress nuclear factor kappa-light-chain-enhancer of activated B cells (NF-κB)-related inflammatory signaling, while estrogen depletion is associated with greater neuroinflammation and oxidative stress.^22,31,32^ Collectively, these mechanisms support our finding that even the low estradiol concentrations, typical of the postmenopausal period, may be associated with lower tau burden and slower tau accumulation.

However, Hu et al.^33^ reported no association between CSF estradiol or estrone and CSF tTau; within the same study, higher CSF estrone was associated with a more favorable Aβ_42_/Aβ_40_ ratio, whereas higher CSF estradiol was associated with higher neuroinflammatory marker levels and smaller regional brain volumes. Importantly, correspondence between peripheral and central sex steroid measures can be incomplete and may vary by analyte and population: early work reported stronger plasma-CSF correlations and suggested that CSF estradiol may track the unbound (free) plasma fraction, consistent with roles of protein binding and blood-brain barrier transport,^34^ whereas more recent mass-spectrometric studies have reported weak serum-CSF correlations for estradiol and other sex steroids.^35^ Assay-validation studies also suggest that CSF estradiol may reflect, in part, local CNS production (e.g., aromatization of precursors) rather than directly mirroring circulating levels.^36^ In this context, Hu et al.’s modest plasma–CSF correlations^33^ and our low collinearity between current HRT use and serum estrogens (**Table S8a** in **Supplement**) support interpreting HRT status, plasma/ serum estrogens, and CSF estrogens as overlapping but non-equivalent indices of estrogenic biology; accordingly, cross-study comparisons should consider which compartment (and assay approach) is used to operationalize estrogen exposure. In our data, current HRT use was only modestly correlated with serum estradiol (r = 0.25, p < 0.001) and estrone (r = 0.35, p < 0.001), and estradiol and estrone were moderately correlated (r = 0.55, p < 0.001). Notably, variance inflation factors indicated minimal multicollinearity among these predictors (VIFs: estrone = 1.53, estradiol = 1.43, HRT current use = 1.15; mean VIF = 1.37), supporting inclusion of current HRT use as a covariate while suggesting that HRT status and measured estrogen concentrations capture related but distinct aspects of estrogen exposure (**Table S8b** in **Supplement**).

The interaction between estradiol and *APOEε4* carrier status in relation to tau biomarkers suggests that genetic risk may shape hormonal associations with AD pathology. Female *APOEε4* carriers have higher AD risk, earlier onset, and faster decline than male carriers,^3,37–39^ and *APOEε4* has been linked to greater tau burden, tau-related neurodegeneration, and a more proinflammatory microglial profile, effects that may indirectly amplify tau pathology beyond its influence on Aβ.^40–43^ Prior work also suggests that estradiol-related sterol pathways may differ by *APOEε4* status, with stronger associations of estradiol with 24(S)-hydroxycholesterol and 27-hydroxycholesterol among carriers^38^ Because 24(S)-hydroxycholesterol is often interpreted as a marker of brain cholesterol turnover, whereas 27-hydroxycholesterol can act as a selective estrogen receptor modulator, these findings raise the possibility that *APOEε4* modifies the coupling between estrogen signaling and cholesterol-derived pathways in postmenopausal women.^38^ In the present study, the cross-sectional association between estradiol and tau was stronger among *APOEε4* carriers, suggesting that *APOEε4*-related differences in lipid metabolism, neuroinflammation, or estrogen signaling may modify the estradiol–tau relationship.

In our study, neither baseline estradiol nor estrone was associated with baseline CSF Aβ_42_ or with changes in Aβ_42_ over time. This contrasts with several lines of preclinical evidence suggesting that estrogen signaling can reduce amyloidogenic processing and promote Aβ clearance through multiple mechanisms, including modulation of amyloid precursor protein (APP) trafficking, downregulation of beta-site amyloid precursor protein-cleaving enzyme 1 (BACE1), enhancement of neprilysin and insulin-degrading enzyme expression, promotion of microglial phagocytosis, and ERβ-related autophagy.^44–46^ Experimental studies have also shown that estrogen deficiency or ovariectomy is associated with greater Aβ accumulation, whereas estradiol replacement can reduce soluble and insoluble Aβ burden.^47–49^ Although experimental studies support estrogen-related modulation of amyloid biology, these mechanisms may not be captured well by baseline peripheral estrogen levels in relation to longitudinal CSF Aβ_42_ in older adults, particularly if estrogen effects on amyloid are timing-specific and concentrated within exposure windows such as the menopausal transition or earlier disease stages.^7,9–11^

In the present study, amyloid positivity was independently associated with steeper longitudinal increases in CSF pTau_181_ and tTau across models, supporting the view that tau accumulation is accelerated downstream of amyloid pathology. This interpretation is consistent with evidence that greater baseline amyloid burden is associated with faster downstream tau accumulation and that this process may be more pronounced in females.^50–54^ However, in models including amyloid-by-estrogen interaction terms, amyloid positivity did not significantly modify the associations of estradiol or estrone with either baseline tau biomarker levels or with changes in these biomarkers over time. Thus, although amyloid positivity identified participants with more rapid tau progression in our cohort, peripheral estrogen levels did not appear to meaningfully alter the strength of the amyloid–tau association. One possible interpretation is that amyloid status captures a major upstream driver of tau progression, whereas circulating estrogens may exert smaller or more context-dependent effects that were difficult to detect in these interaction models.

This study has several strengths. Estradiol and estrone were quantified using LC-MS/MS, an approach well suited to the accurate measurement of low-concentration sex steroids in postmenopausal samples and less susceptible than immunoassays to cross-reactivity and analytic interference ^18,55^. The analysis also leveraged a large, well-characterized EPAD LCS sample with standardized CSF biomarker assessment on the Elecsys platform, providing a strong framework for examining hormone–biomarker associations in a cognitively unimpaired female cohort.

Several limitations should also be considered. Follow-up attrition was substantial and appeared to be informative, as participants with longitudinal data had higher baseline estrogen concentrations and lower baseline Aβ_42_ than those with baseline-only data. In addition, peripheral estrogen concentrations may not adequately reflect central nervous system exposure,^33,56,57^ and although most participants were likely peri- or postmenopausal, non-standardized blood collection timing may still have introduced variability in circulating estrogen measurements.

Finally, HRT exposure was measured only as a binary indicator of current use at baseline, which likely missed important exposure details (e.g., timing, formulation, dose, route, and duration); combined with the low prevalence of HRT use (7.4%), this may have limited power to detect HRT-related effects, warranting cautious interpretation of the null findings.

Future work should integrate peripheral and CSF hormone measurements with comprehensive AD biomarkers to clarify whether circulating estradiol reflects central estrogen activity or whether peripheral and central compartments exert independent effects and should use repeated hormone assessments over time to link within-person hormonal change to biomarker trajectories. Studies with detailed reproductive history and menopausal characteristics, plus detailed HRT exposure (timing, formulation, dose, route, and duration), are needed to separate contemporaneous hormone levels from cumulative and “critical window” effects. Research evaluating estrogen receptor expression, inflammatory markers, and downstream signaling cascades in relation to the APOEε4 variant would provide valuable insights into the biological basis of the observed effect modification.

## Data Availability

EPAD LCS data are available to qualified researchers through a controlled access process via the EPAD/AD Data Initiative data access platform https://ep-ad.org/index.php/open-source-data/.

## ACKNOWLEDGEMENTS

The authors thank the EPAD-LCS participants, without whom this research would not have been possible. The authors also thank Amanda Bannerman for oversight of sample selection and shipment, Kaj Blennow and colleagues at the Clinical Neurochemistry Laboratory, University of Gothenburg, Sweden, for CSF biomarker analyses, and Lee Murphy and the Genetics Core team at the University of Edinburgh for APOE genotyping.

## CONSENT STATEMENT

All participants in each dataset provided informed consent to participate.

