## Supplement for "Associations between serum estradiol and estrone and Alzheimer’s disease biomarkers: an analysis in female participants from the European Prevention of Alzheimer’s Dementia Longitudinal Cohort Study (EPAD LCS)"

### Additional Tables

**Table S1.** Baseline characteristics by follow-up status (attrition analysis)

| Variable | Category | With Follow-up (n=189) M (SD) or % | Dropout (n=677) M (SD) or % | *p* ( t-test/χ2 test) |
|---|---|---|---|---|
| Age (years) | | 65.60 (6.32) | 65.04 (7.45) | 0.35 |
| Education (years) | | 13.79 (3.82) | 14.28 (3.56) | 0.10 |
| Estradiol (log, pg/mL) | | 2.87 (0.60) | 2.28 (1.04) | <.001*** |
| Estrone (log, pg/mL) | | 4.97 (0.46) | 4.83 (0.48) | <.001*** |
| $pTau_{181}$ (pg/mL) | | 19.42 (11.54) | 19.20 (9.76) | 0.80 |
| tTau (pg/mL) | | 223.50 (103.88) | 222.59 (94.31) | 0.91 |
| $A\beta_{42}$ (pg/mL) | | 1338.81 (639.95) | 1466.17 (792.85) | 0.04* |
| $A\beta_{42}$ status | amyloid-positive | 33.33% | 31.17% | 0.57 |
| | amyloid-negative | 66.67% | 68.83% | |
| Ethnicity | Caucasian/White | 86.24% | 71.05% | <.001*** |
| | Hispanic/ Latino | 0% | 1.48% | |
| | Black/Asian/Other | 0% | 1.18% | |
| | Not Reported | 13.76% | 26.29% | |
| *APOE ε4* status | *ε4* non-carrier | 59.79% | 63.37% | 0.37 |
| | *ε4* carrier | 40.21% | 36.63% | |
| HRT current use | Yes | 7.94% | 7.24% | 0.75 |
| | No | 92.06% | 92.76% | |

Notes: * p < .05. ** p < .01. *** p < .001

**Table S2.** Linear mixed model estimates of CSF A$\beta_{42}$ in relation to serum estradiol and estrone (follow-up only re-analysis)

| Predictor | Model 1 | | | | Model 2 | | | |
|---|---|---|---|---|---|---|---|---|
| | Baseline | | Slope | | Baseline | | Slope | |
| | β (SE) | *P* | β (SE) | *P* | β (SE) | *P* | β (SE) | *P* |
| **Estradiol** | | | | | | | | |
| Intercept | 5306.01 (1039.85) | <0.001*** | 139.72 (77.82) | 0.07 | 5337.79 (1050.43) | <0.001*** | 134.69 (82.48) | 0.10 |
| Age at baseline (years) | -60.21 (15.02) | <0.001*** | -1.38 (0.95) | 0.15 | -59.45 (15.09) | <0.001*** | -1.42 (0.96) | 0.14 |
| APOE *ε4* carrier (vs non-carrier) | -337.60 (95.22) | <0.001*** | -8.29 (13.32) | 0.53 | -552.87 (501.57) | 0.27 | 9.08 (55.33) | 0.87 |
| Current HRT use (vs no) | -85.17 (182.48) | 0.64 | 14.25 (25.61) | 0.58 | -92.05 (182.97) | 0.62 | 15.88 (25.79) | 0.54 |
| ln (Estradiol) | 64.77 (87.87) | 0.46 | 5.36 (10.16) | 0.60 | 37.80 (108.33) | 0.73 | 7.70 (13.24) | 0.56 |
| APOE *ε4* carrier × ln (Estradiol) | . | . | . | . | 72.18 (169.38) | 0.67 | -5.30 (18.06) | 0.77 |
| **Estrone** | | | | | | | | |
| Intercept | 3599.02 (1148.95) | 0.002** | 169.16 (99.57) | 0.09 | 3544.94 (1222.84) | 0.004** | 175.06 (117.76) | 0.14 |
| Age at baseline (years) | -59.12 (14.90) | <0.001*** | -2.21 (0.92) | 0.016* | -59.22 (14.92) | <0.001*** | -2.23 (0.93) | 0.016* |
| APOE *ε4* carrier (vs non-carrier) | -319.37 (93.34) | 0.001** | -1.47 (13.16) | 0.91 | -206.32 (1150.77) | 0.86 | -7.34 (132.98) | 0.96 |
| Current HRT use (vs no) | -341.90 (193.19) | 0.08 | 16.28 (27.55) | 0.56 | -341.36 (193.57) | 0.08 | 15.57 (28.08) | 0.58 |
| ln (Estrone) | 375.40 (120.06) | 0.002** | 7.50 (13.78) | 0.59 | 387.57 (152.70) | 0.011* | 6.53 (17.56) | 0.71 |
| APOE *ε4* carrier × ln (Estrone) | . | . | . | . | -22.71 (232.77) | 0.92 | 1.12 (26.44) | 0.97 |

Notes: * p < .05. ** p < .01. *** p < .001

**Table S3.** Linear mixed model estimates of CSF $pTau_{181}$ in relation to serum estradiol (follow-up only re-analysis)

| Predictor | Model 1 | | | | Model 2 | | | | Model 3 | | | |
|---|---|---|---|---|---|---|---|---|---|---|---|---|
| | Baseline | | Slope | | Baseline | | Slope | | Baseline | | Slope | |
| | β (SE) | *P* | β (SE) | *P* | β (SE) | *P* | β (SE) | *P* | β (SE) | *P* | β (SE) | *P* |
| Intercept | 19.81 (11.83) | 0.09 | 0.81 (1.24) | 0.51 | 19.03 (11.93) | 0.11 | 0.06 (1.31) | 0.96 | 19.08 (11.99) | 0.11 | 0.58 (1.24) | 0.64 |
| Age at baseline (years) | 0.02 (0.16) | 0.91 | 0.00 (0.02) | 0.90 | 0.04 (0.16) | 0.78 | 0.00 (0.02) | 0.95 | -0.01 (0.16) | 0.97 | 0.00 (0.02) | 0.98 |
| APOE *ε4* carrier (vs non-carrier) | 1.24 (1.42) | 0.39 | 0.65 (0.20) | 0.001** | -0.92 (7.73) | 0.91 | 2.37 (0.93) | 0.011* | 1.36 (1.42) | 0.34 | 0.67 (0.20) | 0.001** |
| Current HRT use (vs no) | -0.35 (2.71) | 0.90 | 0.59 (0.38) | 0.12 | -0.55 (2.72) | 0.84 | 0.61 (0.38) | 0.11 | -0.80 (2.73) | 0.77 | 0.51 (0.38) | 0.18 |
| Amyloid-positive (vs negative) | 0.37 (0.65) | 0.57 | 0.07 (0.10) | 0.47 | 0.38 (0.64) | 0.56 | 0.08 (0.10) | 0.45 | 7.47 (6.61) | 0.26 | 1.08 (0.65) | 0.10 |
| ln (Estradiol) | -0.91 (1.36) | 0.50 | -0.29 (0.17) | 0.09 | -1.18 (1.67) | 0.48 | -0.01 (0.22) | 0.95 | -0.11 (1.58) | 0.95 | -0.18 (0.18) | 0.34 |
| APOE *ε4* carrier × ln (Estradiol) | . | . | . | . | 0.62 (2.63) | 0.82 | -0.58 (0.31) | 0.06 | . | . | . | . |
| Amyloid-positive × ln (Estradiol) | . | . | . | . | . | . | . | . | -2.50 (2.26) | 0.27 | -0.33 (0.21) | 0.11 |

Notes: * p < .05. ** p < .01. *** p < .001

**Table S4.** Linear mixed model estimates of CSF tTau in relation to serum estradiol (follow-up only re-analysis)

| Predictor | Model 1 | | | | Model 2 | | | | Model 3 | | | |
|---|---|---|---|---|---|---|---|---|---|---|---|---|
| | Baseline | | Slope | | Baseline | | Slope | | Baseline | | Slope | |
| | β (SE) | *P* | β (SE) | *P* | β (SE) | *P* | β (SE) | *P* | β (SE) | *P* | β (SE) | *P* |
| Intercept | 193.22 (121.67) | 0.11 | 11.91 (9.36) | 0.20 | 182.61 (123.43) | 0.14 | 10.03 (9.75) | 0.30 | 179.36 (122.55) | 0.14 | 8.88 (9.46) | 0.35 |
| Age at baseline (years) | 0.55 (1.71) | 0.75 | -0.07 (0.12) | 0.55 | 0.64 (1.71) | 0.71 | -0.07 (0.12) | 0.57 | 0.28 (1.70) | 0.87 | -0.08 (0.12) | 0.49 |
| APOE *ε4* carrier (vs non-carrier) | 20.55 (13.39) | 0.13 | 5.72 (1.59) | <0.001*** | 32.29 (67.59) | 0.63 | 9.90 (6.67) | 0.14 | 22.00 (13.28) | 0.10 | 6.00 (1.60) | <0.001*** |
| Current HRT use (vs no) | 4.98 (25.70) | 0.85 | 4.87 (3.14) | 0.12 | 4.36 (25.69) | 0.87 | 5.06 (3.17) | 0.11 | -1.22 (25.71) | 0.96 | 3.86 (3.19) | 0.23 |
| Amyloid-positive (vs negative) | -5.74 (7.09) | 0.42 | 2.50 (1.01) | 0.014* | -5.65 (7.09) | 0.43 | 2.45 (1.02) | 0.016* | 94.56 (63.36) | 0.14 | 13.82 (6.22) | 0.026* |
| ln (Estradiol) | -4.75 (11.99) | 0.69 | -2.30 (1.19) | 0.05 | -2.94 (14.89) | 0.84 | -1.75 (1.48) | 0.24 | 5.92 (14.12) | 0.68 | -0.97 (1.41) | 0.49 |
| APOE *ε4* carrier × ln (Estradiol) | . | . | . | . | -4.40 (23.16) | 0.85 | -1.35 (2.15) | 0.53 | . | . | . | . |
| Amyloid-positive × ln (Estradiol) | . | . | . | . | . | . | . | . | -35.09 (21.81) | 0.11 | -3.83 (2.01) | 0.06 |

Notes: * p < .05. ** p < .01. *** p < .001

**Table S5.** Linear mixed model estimates of CSF $pTau_{181}$ in relation to serum estrone (follow-up only re-analysis)

| Predictor | Model 1 | | | | Model 2 | | | | Model 3 | | | |
|---|---|---|---|---|---|---|---|---|---|---|---|---|
| | Baseline | | Slope | | Baseline | | Slope | | Baseline | | Slope | |
| | β (SE) | *P* | β (SE) | *P* | β (SE) | *P* | β (SE) | *P* | β (SE) | *P* | β (SE) | *P* |
| Intercept | 17.67 (14.12) | 0.21 | 2.36 (1.58) | 0.13 | 9.01 (15.30) | 0.56 | 1.74 (1.80) | 0.34 | 15.47 (14.38) | 0.28 | 2.04 (1.56) | 0.19 |
| Age at baseline (years) | 0.07 (0.16) | 0.66 | 0.00 (0.01) | 0.82 | 0.05 (0.16) | 0.78 | 0.00 (0.01) | 0.87 | 0.05 (0.16) | 0.74 | 0.00 (0.01) | 0.98 |
| APOE *ε4* carrier (vs non-carrier) | 0.95 (1.43) | 0.51 | 0.61 (0.20) | 0.002** | 27.56 (17.07) | 0.11 | 2.80 (2.00) | 0.16 | 0.97 (1.43) | 0.50 | 0.62 (0.20) | 0.002** |
| Current HRT use (vs no) | -0.42 (2.93) | 0.89 | 0.84 (0.41) | 0.039* | -0.67 (2.92) | 0.82 | 0.80 (0.40) | 0.046* | -0.55 (2.92) | 0.85 | 0.77 (0.40) | 0.06 |
| Amyloid-positive (vs negative) | 0.34 (0.65) | 0.60 | 0.07 (0.10) | 0.52 | 0.32 (0.65) | 0.62 | 0.07 (0.10) | 0.51 | 17.84 (16.87) | 0.29 | 2.51 (1.50) | 0.09 |
| ln (Estrone) | -0.75 (1.87) | 0.69 | -0.50 (0.22) | 0.026* | 1.32 (2.38) | 0.58 | -0.36 (0.29) | 0.21 | -0.03 (1.97) | 0.99 | -0.40 (0.23) | 0.09 |
| APOE *ε4* carrier × ln (Estrone) | . | . | . | . | -5.39 (3.45) | 0.12 | -0.44 (0.40) | 0.27 | . | . | . | . |
| Amyloid-positive × ln (Estrone) | . | . | . | . | . | . | . | . | -3.63 (3.46) | 0.29 | -0.49 (0.30) | 0.10 |

Notes: * $p < .05$. ** $p < .01$. *** $p < .001$

**Table S6**. Linear mixed model estimates of CSF tTau in relation to serum estrone (follow-up only re-analysis)

| Predictor | Model 1 | | | | Model 2 | | | | Model 3 | | | |
|---|---|---|---|---|---|---|---|---|---|---|---|---|
| | Baseline | | Slope | | Baseline | | Slope | | Baseline | | Slope | |
| | β (SE) | *P* | β (SE) | *P* | β (SE) | *P* | β (SE) | *P* | β (SE) | *P* | β (SE) | *P* |
| Intercept | 153.03 (140.92) | 0.28 | 19.79 (13.00) | 0.13 | 83.78 (150.06) | 0.58 | 11.10 (14.54) | 0.45 | 108.20 (142.81) | 0.45 | 14.06 (13.15) | 0.29 |
| Age at baseline (years) | 1.10 (1.69) | 0.52 | -0.07 (0.12) | 0.57 | 0.94 (1.69) | 0.58 | -0.07 (0.12) | 0.57 | 0.86 (1.68) | 0.61 | -0.10 (0.12) | 0.41 |
| APOE *ε4* carrier (vs non-carrier) | 19.30 (13.49) | 0.15 | 5.15 (1.60) | 0.001** | 269.91 (163.93) | 0.10 | 32.66 (16.83) | 0.05 | 19.16 (13.38) | 0.15 | 5.28 (1.61) | 0.001** |
| Current HRT use (vs no) | 1.27 (28.01) | 0.96 | 6.51 (3.45) | 0.06 | -1.56 (27.79) | 0.96 | 6.48 (3.54) | 0.07 | -1.60 (27.82) | 0.95 | 5.54 (3.51) | 0.12 |
| Amyloid-positive (vs negative) | -5.58 (7.07) | 0.43 | 2.19 (1.00) | 0.028* | -5.71 (7.06) | 0.42 | 2.16 (1.01) | 0.032* | 294.51 (165.45) | 0.08 | 36.33 (15.12) | 0.016* |
| ln (Estrone) | -1.37 (17.61) | 0.94 | -2.94 (1.80) | 0.10 | 14.57 (21.73) | 0.50 | -1.20 (2.14) | 0.58 | 11.03 (18.57) | 0.55 | -1.37 (1.96) | 0.49 |
| APOE *ε4* carrier × ln (Estrone) | . | . | . | . | -51.04 (33.18) | 0.12 | -5.52 (3.34) | 0.10 | . | . | . | . |
| Amyloid-positive × ln (Estrone) | . | . | . | . | . | . | . | . | -61.83 (33.88) | 0.07 | -6.89 (3.03) | 0.023* |

Notes: * p < .05. ** p < .01. *** p < .001

**Table S7**. Stratified analysis by APOE *ε4* carrier status: Linear mixed model estimates of tau in relation to serum estradiol & estrone

| Predictor | pTau181 | | | | | | | | tTau | | | | | | | |
|---|---|---|---|---|---|---|---|---|---|---|---|---|---|---|---|---|
| | APOE *ε4* carriers | | | | APOE *ε4* non-carriers | | | | APOE *ε4* carriers | | | | APOE *ε4* non-carriers | | | |
| | Baseline | | Slope | | Baseline | | Slope | | Baseline | | Slope | | Baseline | | Slope | |
| | β (SE) | p | β (SE) | p | β (SE) | p | β (SE) | p | β (SE) | p | β (SE) | p | β (SE) | p | β (SE) | p |
| **Estradiol** | | | | | | | | | | | | | | | | |
| Intercept | 25.82 (14.83) | 0.08 | -1.38 (0.85) | 0.10 | 11.45 (10.61) | 0.28 | 0.45 (0.40) | 0.26 | 322.31 (161.87) | 0.046* | -8.83 (8.26) | 0.29 | 135.80 (114.46) | 0.24 | 4.65 (4.13) | 0.26 |
| Age at baseline (years) | -0.03 (0.22) | 0.88 | 0.04 (0.01) | 0.002** | 0.11 (0.16) | 0.49 | -0.00 (0.01) | 0.64 | -0.77 (2.44) | 0.75 | 0.29 (0.11) | 0.011* | 1.32 (1.73) | 0.45 | -0.03 (0.05) | 0.59 |
| Current HRT use (vs no) | 2.18 (2.09) | 0.30 | 0.01 (0.29) | 0.96 | 0.54 (1.47) | 0.71 | 0.02 (0.17) | 0.89 | 23.14 (19.66) | 0.24 | 0.78 (2.78) | 0.78 | 7.69 (14.67) | 0.60 | 0.61 (1.80) | 0.73 |
| Amyloid-positive ( vs negative) | 0.10 (0.72) | 0.89 | 0.17 (0.13) | 0.20 | 0.07 (0.70) | 0.92 | 0.28 (0.07) | <0.001*** | -5.36 (7.65) | 0.48 | 1.93 (1.36) | 0.16 | -15.69 (7.10) | 0.027* | 2.70 (0.77) | <0.001*** |
| ln (Estradiol) | -1.69 (0.63) | 0.01 | -0.10 (0.08) | 0.20 | -0.31 (0.38) | 0.41 | -0.06 (0.04) | 0.13 | -17.96 (5.91) | 0.002** | -1.13 (0.74) | 0.13 | -2.73 (3.76) | 0.47 | -0.53 (0.40) | 0.18 |
| **Estrone** | | | | | | | | | | | | | | | | |
| Intercept | 31.96 (16.42) | 0.05 | -1.06 (1.20) | 0.38 | 9.99 (10.50) | 0.34 | 1.14 (0.61) | 0.06 | 329.37 (175.30) | 0.06 | -8.68 (11.66) | 0.46 | 91.76 (118.05) | 0.44 | 10.25 (6.20) | 0.10 |
| Age at baseline (years) | -0.02 (0.22) | 0.92 | 0.04 (0.01) | 0.002** | 0.12 (0.15) | 0.43 | -0.01 (0.01) | 0.12 | -0.35 (2.44) | 0.88 | 0.31 (0.12) | 0.008** | 1.59 (1.69) | 0.35 | -0.09 (0.05) | 0.11 |
| Current HRT use (vs no) | 1.67 (2.15) | 0.44 | -0.04 (0.30) | 0.88 | 0.07 (1.48) | 0.96 | 0.14 (0.21) | 0.49 | 12.68 (20.33) | 0.53 | -0.22 (2.85) | 0.94 | 0.95 (14.65) | 0.95 | 0.95 (2.14) | 0.66 |
| Amyloid-positive (vs negative) | 0.04 (0.72) | 0.95 | 0.17 (0.13) | 0.20 | 0.53 (0.68) | 0.44 | 0.25 (0.08) | 0.001** | -5.72 (7.69) | 0.46 | 1.94 (1.37) | 0.16 | -12.65 (7.04) | 0.07 | 2.75 (0.82) | 0.001** |
| ln (Estrone) | -2.22 (1.32) | 0.09 | -0.13 (0.16) | 0.43 | 0.11 (0.77) | 0.89 | -0.09 (0.09) | 0.30 | -15.70 (12.55) | 0.21 | -0.88 (1.57) | 0.58 | 4.69 (7.59) | 0.54 | -0.72 (0.92) | 0.43 |

Notes: * p < .05. ** p < .01. *** p < .001

**Table S8**. Correlations and multicollinearity diagnostics for current hormone replacement therapy (HRT) use and log-transformed estradiol and estrone

a. Pearson correlations among current HRT use and log-transformed estradiol and estrone

| | HRT current use | Estradiol (log) | Estrone (log) |
|---|---|---|---|
| HRT current use | 1 | | |
| Estradiol (log) | 0.25*** | 1 | |
| Estrone (log) | 0.35*** | 0.55*** | 1 |

Notes: * p < .05. ** p < .01. *** p < .001

b. Variance inflation factors (VIFs) for current HRT use and log-transformed estradiol and estrone

| Variable | VIF | 1/VIF |
|---|---|---|
| HRT current use | 1.15 | 0.87 |
| Estradiol (log) | 1.43 | 0.7 |
| Estrone (log) | 1.53 | 0.65 |
| Mean VIF | 1.37 | |
